# Introducing Infrastructure as Code for deployment and governance of medical AI: A case study

**DOI:** 10.64898/2026.10.12.26365327

**Authors:** Markus Vogel, Nils Reuter, Sebastian Ruf, Ayush Noori, Julian Madrid, Ran Balicer, Martin Witzenrath, Leif Erik Sander, Matthias I. Gröschel

**Affiliations:** Department of Infectious Diseases, Respiratory, and Critical Care Medicine, Charité University Medical Center Berlin, Berlin, Germany; Microsoft GmbH, Munich, Germany; Department of Engineering Science, University of Oxford, Oxford, United Kingdom; Department of Biomedical Informatics, Harvard Medical School, Boston, Massachusetts, United States of America; Institute for Artificial Intelligence in Medicine (IKIM), Charité University Medical Center Berlin, Germany; Clalit Research Institute, Innovation Division, Clalit Health Services, Ramat Gan, Israel; School of Public Health, Faculty of Health Sciences, Ben Gurion University of the Negev, Be’er Sheva, Israel; Berlin Institute of Health, Berlin, Germany

**Keywords:** artificial intelligence, healthcare, medical documents, inconsistencies, large language models, infrastructure as code

## Abstract

Clinical AI deployment rarely reaches the bedside because both workstation integration and governed backend provisioning are challenging. We introduce a workflow that allows rapid and governed access to AI functionalities through a browser interface or API, enabling privacy-compliant data analysis. The complex setup for compliant healthcare applications such as a secure backend, private AI model endpoint, and managed identity configuration is encoded by itself, a concept known as Infrastructure as Code *(IaC)*. IaC is a concept in which networks, access rights, and service endpoints are written down as machine-readable files that can be reviewed once and executed identically in any institution. Here, we apply this concept to a clinical use case. The process of deploying a governed, private clinical AI solution using this approach in a tertiary care center took, on average, 310.3 ± 196.6 seconds *(n=25, mean ± SD)*. To further demonstrate the clinical value of this approach, we developed and deployed an LLM-based inconsistency checker, which identifies textual inconsistency in medical documents. When evaluating the tool on a set of 1,347 physician-approved clinical documents, the tool flagged a clinically severe inconsistency in 105 clinical documents *(7*.*8%, approximately 1 in 13)*. In a severity-stratified physician audit, 149 of 150 sampled findings were confirmed *(population-weighted PPV 98*.*2%; CI 94*.*8 - 100*.*0)*. In summary, we present the application of IaC frameworks to a healthcare use case and demonstrate that clinically useful AI applications can be deployed to physician workstations in hospital-governed cloud environments within a few minutes.

**Description:** To overcome the implementation barriers for use of AI in healthcare, we adapted Infrastructure as Code to a clinical use case and deployed a hospital-governed large language model service that clinicians reach from a browser without workstation software. Provisioned by Infrastructure as Code the service became available in 310 seconds on average, and its first clinical application, an inconsistency checker, detected inconsistencies in clinical documents with high positive predictive value.

## Introduction

Many healthcare practitioners have encountered the same sequence: a promising AI tool is demonstrated but fails to be implemented. Despite the rapid pace of innovation around medical artificial intelligence (AI) and large language models (LLMs) in drafting and assessing hospital discharge documentation,^1^ their transition into clinical practice remains limited. This ‘implementation gap’ reflects opportunity costs, fragmented data environments, governance requirements, and limited workflow integration.^2,3^

Today, clinical AI infrastructure typically involves network segments, identity boundaries, data-protection reviews, and software deployments that must be assembled by hand at every institution that wants to deploy it. This manual approach creates large administrative, regulatory and IT overheads for every new data-driven tool practitioners wish to implement.^4,5^ More than a decade ago, other sectors solved this problem through a paradigm known as Infrastructure as Code (IaC). Instead of manually configuring cloud environments, IT teams write the infrastructure requirements into machine-readable files that can be reviewed, versioned, and executed identically anywhere. For example, a company can use IaC to deploy several identical servers by running a single script. If security updates are required, engineers update the code once for roll-out across all deployed servers, eliminating human error and saving significant work hours.^6,7^ Despite these benefits and widespread success in fields like finance and manufacturing, healthcare has seen only isolated uptake of IaC. A recent study proposed and piloted a cloud-based IaC architecture for federated learning in dementia imaging,^5^ but codified deployment of a governed clinical language-model service, and the time this takes, have not been reported, although similar logics have long been embraced for clinical care itself. For example, written, reproducible protocols are widely used in medicine to standardize treatments and have helped reduce patient harm.^8^

Once the requirements are defined and approved by a data protection officer, the codified infrastructure can be re-executed unchanged in the next department or institution. All processing then occurs within the hospital’s own IaC-defined cloud environment, so patient data never leave hospital governance and are not used for model training.

This IaC framework for use in hospitals implements a governed Large Language Model (LLM) service that clinicians can invoke without installing any workstation software. We evaluated an inconsistency detection service as its specific clinical use case to map inconsistencies in clinical documents at scale and to streamline hospital quality reporting.^9^

### Setting and Implementation

#### Architecture of the Reproducible AI Infrastructure

We engineered an IaC framework that encapsulates all necessary components for secure clinical AI deployment within a single, version-controlled repository (Table 1).

**Table 1:** Infrastructure as Code governance requirements, codified components, and clinical use. Bicep, Microsoft’s declarative language for Azure resources, compiled to Azure Resource Manager (ARM) template

| Requirement | Codified Component | Clinical Use |
| --- | --- | --- |
| Governed Application Hosting | Infrastructure as Code (Bicep, compiled to ARM), frontend JavaScript, backend Python, Function App | Versioned, reproducible and citable backend and frontend functionality |
| Private Model Access | Virtual Network, Private Endpoint, interchangeable Large Language Model (LLM) isolated from the public internet, EU data zone processing by default | Secure, governed LLM backend, model weights are not trained or changed by clinical use |
| Controlled Data Retention | No persistence of clinical text by the application, required Functions runtime storage with shared key access disabled, telemetry limited to status codes and durations | Single data entry point remains the clinical application |
| Identity Management | Role Based Access Control (RBAC) for all components, codified user authentication via OpenID Connect against Microsoft Entra ID; the built-in authentication layer also supports other OpenID Connect providers | Single sign-on fully supported, no new password required, seamless clinical workflow integration, but still full access control for local IT |

The framework treats the model endpoint as a modular deployment parameter, allowing hospital IT departments to upgrade or substitute underlying models without requiring any architectural modifications to the core application (Figure 1A).

**Figure 1:**
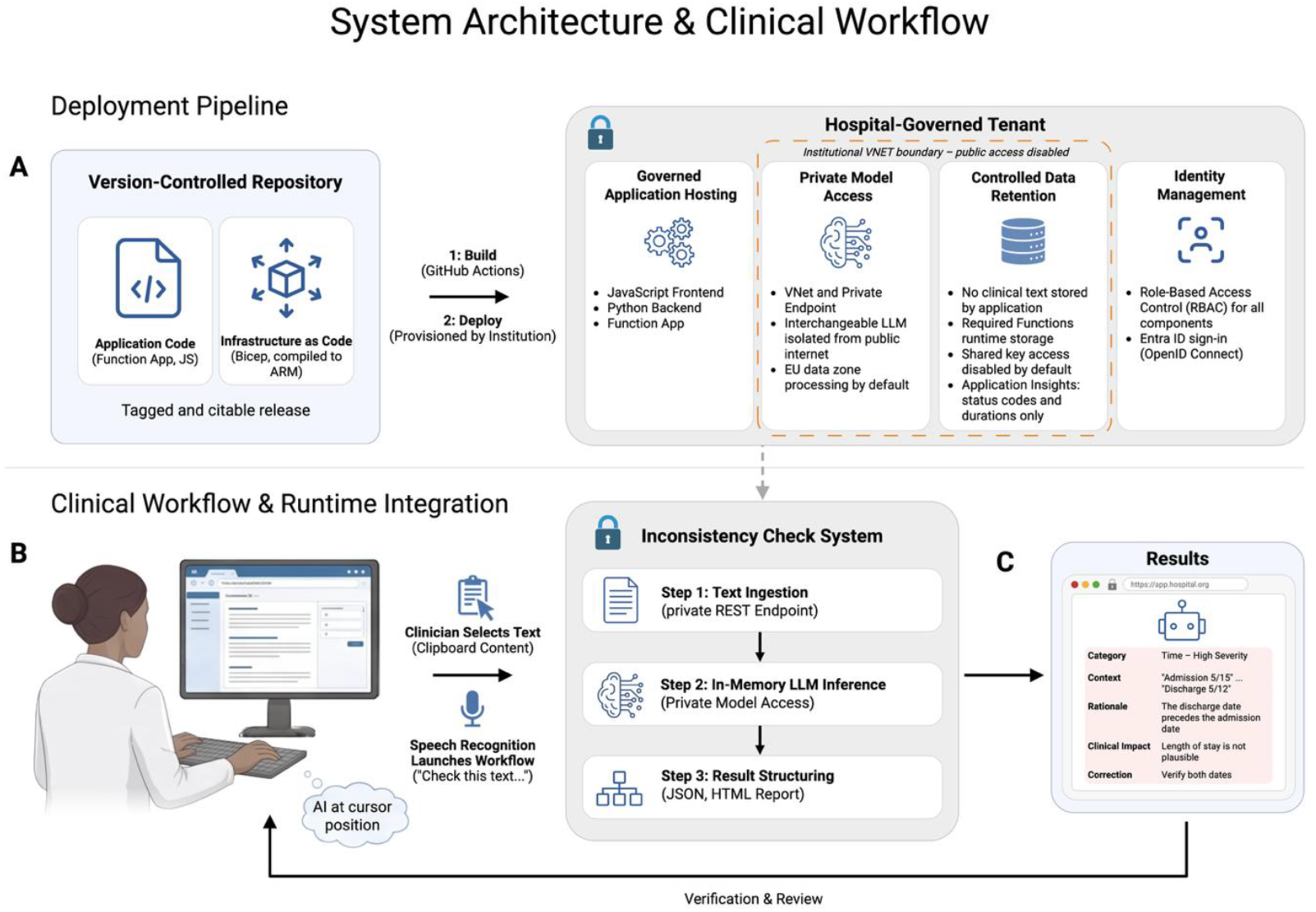
Governed deployment and runtime workflow. (A) The entire system (infrastructure (Bicep), application, and governance) is provided in a single version-controlled repository. (B) At runtime, a clinician submits selected or dictated clinical text through a browser interface without dedicated workstation software. (C) The application processes text in memory within a secure, governed network with private LLM endpoints and returns a structured inconsistency report for physician verification.

#### Integration into clinical workflow

Clinicians can use the system at cursor level. Access is controlled centrally through the hospital’s identity management (Figure 1A). Selected or dictated text is pasted into the browser interface, or the frontend is invoked by speech recognition software (Figure 1B,C).

#### Deployment evaluation

To assess the ease of implementation we timed 25 independent deployments (five models × five attempts in randomized complete blocks, run serially) from initiation of the infrastructure deployment to the first successful model response over the private endpoint. This corresponds to the time a hospital’s IT department would need to provide the clinician with the governed AI tool at bedside.

#### Use case evaluation

To demonstrate the clinical usefulness of this framework, we designed an inconsistency checker to identify inconsistencies in medical documents. We evaluated this tool on de-identified physician-authored clinical documents obtained from routine clinical practice.^10^ Documents were provided as native DOCX files and stored in a local directory reflecting standard hospital export workflows, and results were returned in structured form. Evaluation took place in two phases (Table 2).

**Table 2:** Study design and cohorts. PPV, positive predictive value. *Full panel: Claude Opus 4.7, GPT-5.5, Mistral Large 3, DeepSeek V3.2, and GPT-5.4-nano. Development phase and validation phase comprise disjoint documents (total corpus 1,447 documents)

| Characteristic | Development phase (100 letters) | Validation phase (1,347 letters) |
| --- | --- | --- |
| Purpose | Concept validation (human ground truth) | Scale validation (precision audit) |
| Documents, n | 100 | 1,347 |
| Patients | 100 | Not established (de-identified; no patient identifiers) |
| Cohort period | 2025 | 2025 (date-shifted for de-identification) |
| Setting | Two wards, Department of Infectious Diseases, Respiratory, and Critical Care Medicine, Charité - Universitätsmedizin Berlin | Two wards, Department of Infectious Diseases, Respiratory, and Critical Care Medicine, Charité - Universitätsmedizin Berlin |
| LLM (version) | GPT-4o via browser interface | Five-model panel via governed API; reference model Claude Opus 4.7* |
| Prompt | Baseline prompt (Supplementary Table 1) | Refined prompt (Supplementary Table 1) |
| Reviewer | 4 senior physicians (consensus) | 1 senior physician, 2 for severe findings |
| Review method | Full adjudication of all 576 flagged items | Severity-stratified random subsample: 150 findings (55 severe, 60 relevant, 35 minor) plus 15 model-negative letters; fixed seed |
| Primary endpoints | PPV; severity agreement; category, explanation, and correction quality | Per-stratum PPV; severity appropriateness; correction adoptability |

In the development phase (100 documents), we used a system prompt that defined the model’s role as a senior physician correcting the letter and asked it to apply logical reasoning to flag obviously erroneous content. Each finding was rated from 1 (minor ambiguity without consequence) to 9 (must be corrected); output was a table. For analysis, scores 7-9 were labelled severe, 4-6 relevant, and 1-3 minor in both phases. Four senior physicians reviewed every inconsistency flagged across all 100 clinical documents and recorded one consensus judgment per finding (true or false positive).

In the validation phase (1,347 documents), we restricted the prompt to internal logical contradictions not requiring medical knowledge, defined severity bands, instructions to be conservative, and JSON output (Supplementary Table S1). Because exhaustive review of all inconsistencies detected in 1,347 letters across five models was infeasible, human adjudication was restricted to a fixed-seed, severity-stratified sample from the reference index model selected for physician audit (Claude Opus 4.7). The sample was adjudicated by one senior physician; in an additional check, a second senior physician independently reviewed the severe findings. Each flagged issue was assessed for whether it was a genuine inconsistency, whether its assigned severity was appropriate, and whether the proposed correction was clinically adoptable. Sensitivity and clinical outcomes were not assessed (Table 2). Claude Opus 4.7 was selected as the index model for physician audit, because it generated the highest number of candidate findings, provided the broadest category coverage, and produced fully schema-conformant output. This high-yield selection created a large and heterogeneous pool for severity-stratified sampling and was not intended to indicate superior model accuracy.Table 2

#### Statistical methods

Data were analyzed using descriptive statistics: continuous variables are reported as mean (SD) or median (IQR), and categorical variables as counts and percentages.

Positive predictive values (PPVs) and conditional quality proportions are presented with 95% Wilson score confidence intervals, as some proportions were close to 0% or 100%. The overall PPV was computed as the weighted average of stratum-specific PPVs, using the index model’s observed flag distribution (1.6% severe, 36.2% relevant, 62.1% minor) as weights. No hypothesis testing or formal comparisons between models or phases were performed, as the study was designed as a precision audit rather than a comparative evaluation.

#### Ethics

The study was approved by the institutional review board of Charité - Universitätsmedizin Berlin (EA2/322/23).

## Results

### Deployment

Across 25 deployments spanning five models (five deployments per model), the median time from infrastructure deployment initiation to the first valid model response was 237.9 s (range 150.7-953.1 seconds); the mean was 310.3 ± 196.6 seconds (mean ± SD) (Figure 2A).

**Figure 2:**
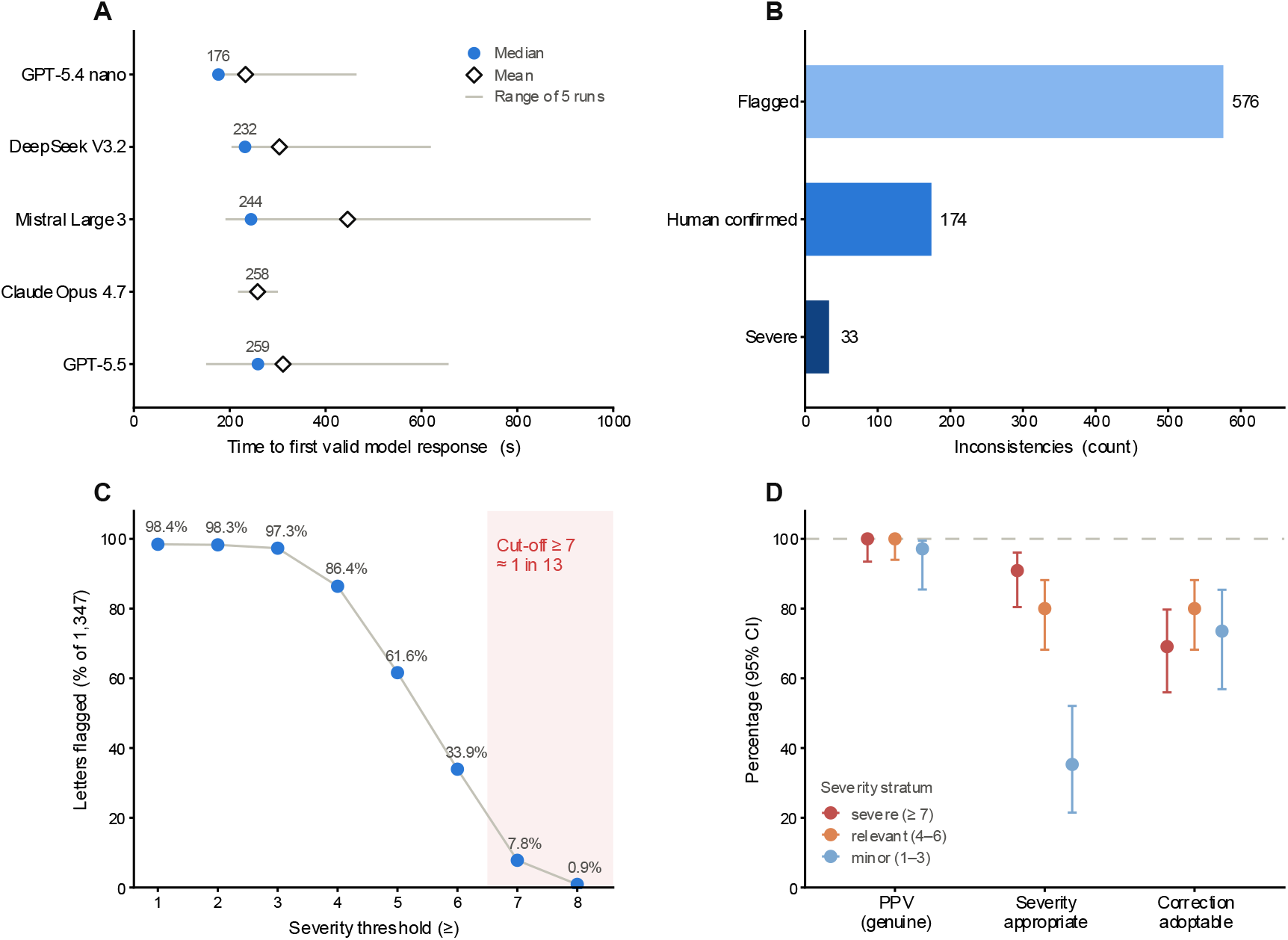
Deployment and Validation. (A) Deployment latency across five frontier models (5 runs each): overall, median time to first valid response was 237.9 seconds (range 150.7 seconds (GPT-5.5) - 953.1 s (Mistral Large 3), n=25). Circles, median; diamonds, mean; grey lines, range. (B) Development phase (100 letters, GPT-4o baseline): 576 flagged: 174 human-confirmed genuine (30%), 33 rated severe by both LLM and physician, illustrating over-alerting. (C) Validation phase (1,347 letters): 98.4% of letters carried ≥1 flag, but severity acts as a tunable filter: raising the threshold to ‘severe’ (≥7 on the 1-9 scale) yields a review queue of 105 letters (7.8%, approximately 1 in 13). (D) Severity-stratified physician audit: positive predictive value (PPV) is near-complete across strata, whereas severity-appropriateness falls for minor findings and correction adoptability is intermediate: detection is reliable, calibration needs the clinician. Points, proportion; error bars, 95% Wilson CIs.

### Study population

A total of 1,447 de-identified, clinician-authored clinical documents from 2025 were collected from the Departments of Infectious Diseases, Respiratory, and Critical Care Medicine at Charité University Medical Center Berlin and split into a development set (n = 100) and a validation set (n = 1,347). All documents had already undergone routine review and sign-off by at least two physicians. The development cohort (n = 100) was majority male (60%, n = 60), with a median age of 63 years (IQR = 26 years) and a median hospital stay of 5 days (IQR = 8 days). The most common primary diagnoses were community-acquired pneumonia (20%), non-small cell lung cancer (9%), and chronic obstructive pulmonary disease (7%). The validation phase cohort (n = 1,347)was comparable in age and length of stay (median age 63 years (IQR = 20 years) with a median stay of 6 days (IQR = 8 days).

### Detection performance

#### Development phase

In development phase (100 letters), the LLM identified a total of 576 potential inconsistencies (5.76 (SD = 1.15) per document), with content-related issues constituting the majority (369 instances, 64%), followed by temporal inconsistencies (n = 87, 15%), structural issues (n = 69, 12%), linguistic inconsistencies (n = 46, 8%), and other inconsistencies (n = 5, 1%). The tool rated 17 (3%) of all 576 flagged inconsistencies as minor, 224 (39%) as relevant, and 335 (58%) as severe.

Of the 576 flagged inconsistencies, 174 (30%) were human confirmed as justified (1.74 per document, SD = 1.63), whereas the remaining 402 (70%) were not confirmed (Figure 2B). No factual hallucinations were observed. The non-confirmed flags predominantly reflected the model reporting apparently missing information that in fact requires implicit institutional knowledge unavailable from the document text (for example, the routine timing and location of a scheduled follow-up appointment).

Among the 174 confirmed inconsistencies, the LLM assigned the correct category in 156 (90%), provided a reasonable explanation in 168 (97%), and proposed a correction that could be adopted without further effort in 147 (84%). Severity was judged appropriate in 66 (38%). While the category was considered incorrect in 18 (18/174, 10%), the proposed correction was inadequate in 27 (27/174, 16%) and the explanation unjustified in 6 (6/174, 3%). Overall, of 335 findings the LLM rated severe, only 33 (9.9%) were also rated severe on physician review, indicating poor alignment. Examples of severe inconsistencies include medication dosage contradictions, conflicting tumor histology, and missed pathogen resistances.

#### Validation phase

Guided by the development phase error analysis, we refined the system prompt (Supplementary Table S1) and applied it unchanged across a five-model panel. All five models flagged an inconsistency in nearly every letter (1,345/1,347 flagged by at least one model; 1,171 (87%) by all five), agreeing far more on which letters contained inconsistencies than on how many (Supplementary Table S2). Compared with the development phase, the PPV over all flags rose from 30.2% (174/576) to 98.2% (weighted) and the share of findings rated severe fell from 58% to 1.6%, while the number of flags per document changed little (5.76 vs. 4.91). Restricted to severe findings, the reference model flagged 105 of 1,347 letters (7.8%) showing a review queue of roughly 1 in 13 clinical documents (Figure 2C). In the stratified audit, PPV was 100% for severe (55/55, confirmed by second reviewer) and relevant (60/60) and 97.1% for minor findings (34/35). Among confirmed findings, severity was found appropriate in 90.9%, 80.0%, and 35.3%, and corrections were adoptable in 69.1%, 80.0%, and 73.5% of severe, relevant, and minor findings, respectively (Table 3, Figure 2D). In the 15 model-negative letters, the reviewing physician found no inconsistencies (0/15).

**Table 3:** Clinical validation results. PPV, positive predictive value; CI, confidence interval (Wilson). Development phase (100 letters) quality metrics are conditional on the 174 confirmed inconsistencies. Validation phase (1,347 letters) severity and correction metrics are conditional on confirmed findings within each stratum. *Population-weighted by the reference model’s severity-band prevalence (severe 1.6%, relevant 36.2%, minor 62.1%). 149 of the 150 reviewed findings were confirmed genuine (the single false positive was a minor-severity finding); because severe findings were deliberately oversampled, the overall value is weighted giving 98.2%; design-based 95% CI. Phases used different models, prompts, and sampling designs and are not a direct head-to-head comparison

| Metric | Scope | Events / total | Value (95% CI) |
| --- | --- | --- | --- |
| **Panel A - Development phase (100 letters): 100 letters, GPT-4o via browser (full adjudication)** |  |  |  |
| Flagged inconsistencies | all letters | 576 | 5.76 per letter |
| Confirmed genuine (PPV) | all flags | 174/576 | 30.2% (26.6-34.1) |
| Severity appropriate | confirmed | 66/174 | 37.9% (31.1-45.3) |
| Category correct | confirmed | 156/174 | 89.7% (84.2-93.4) |
| Explanation reasonable | confirmed | 168/174 | 96.6% (92.7-98.4) |
| Correction adoptable | confirmed | 147/174 | 84.5% (78.4-89.1) |
| Severe by LLM and physician | LLM rated severe | 33/335 | 9.9% (7.1-13.5) |
| **Panel B - Validation phase (1,347 letters): 1,347 letters, Claude Opus 4.7** |  |  |  |

Table 3: Clinical validation results.
| *Detection yield (all 1,347 letters)* |  |  |  |
| --- | --- | --- | --- |
| Flagged inconsistencies | all letters | 6,613 | 4.91 per letter |
| Severity of flags<br>(minor/relevant/severe) | all flags | 4,109/2,396/108 | 62.1% / 36.2% / 1.6% |
| *Validation - severity-stratified sub-sample* |  |  |  |
| Detection (PPV) | severe | 55/55 | 100.0% (93.5-100.0) |
| Detection (PPV) | relevant | 60/60 | 100.0% (94.0-100.0) |
| Detection (PPV) | minor | 34/35 | 97.1% (85.5-99.5) |
| Detection (PPV) | overall, weighted* | - | 98.2% (94.8-100.0) |
| Severity appropriate | severe (confirmed) | 50/55 | 90.9% (80.4-96.1) |
| Severity appropriate | relevant (confirmed) | 48/60 | 80.0% (68.2-88.2) |
| Severity appropriate | minor (confirmed) | 12/34 | 35.3% (21.5-52.1) |
| Correction adoptable | severe (confirmed) | 38/55 | 69.1% (56.0-79.7) |
| Correction adoptable | relevant (confirmed) | 48/60 | 80.0% (68.2-88.2) |
| Correction adoptable | minor (confirmed) | 25/34 | 73.5% (56.9-85.4) |

## Discussion

To overcome implementation barriers to bringing clinical AI to everyday healthcare workstations, we established a codified implementation framework designed for deployment within hospital governance. Our proposed architecture made a fast and governed deployment feasible, and the tool was available to the clinicians within minutes. The presented deployment architecture, governed through Infrastructure as Code, demonstrates that security and compliance requirements can be encoded, removing a principal barrier to clinical AI adoption.

Even among documents that had passed standard dual-physician review, the inconsistency check surfaced a clinically severe inconsistency in approximately 1 in 13 documents (7.8%) with severity endorsed by the reviewing physician in 90.9% of audited severe findings. In the development phase, 70% of flags were not confirmed, a rate that as a consequence would risk ‘over-alerting’ users, possibly leading to alert fatigue.^11,12^ The refined prompt and model took over the triage that the physician had performed manually, and the severity threshold that reduces alert burden also determines how many letters enter the review queue. Calibration was weakest in the lowest category: severity was judged appropriate for only 12 of 34 confirmed minor findings, so findings below the review threshold may still need attention. The substantial number of inconsistencies that escaped prior human revision underlines the need for improvements in the clinical workflow.

Nevertheless, this study has several limitations. First, adjudication relied on senior physician judgment without a standardized criterion-based catalogue, Development-phase judgments of four physicians were recorded as consensus without individual ratings, and only the 55 severe validation findings were double-reviewed (55/55 agreement). Interrater reliability could therefore not be quantified. The results should be read as high-precision detection rather than a definitive reference standard. Second, sensitivity and specificity were not measured; 15 model-negative letters are too few to estimate missed inconsistencies. Third, the five-model results were descriptive; no formal accuracy-versus-cost comparison was performed. Finally, the evaluation was conducted in one clinical department.

Because the environment is easily reproducible by construction, the same deployment can serve as the basis for further prospective, multi-center studies of clinical AI.

## Data Availability

All data produced in the present study are available upon reasonable request to the authors

## Funding

This study received no external funding.

## Data Sharing

Patient-level clinical documents cannot be shared publicly because of privacy and ethics restrictions. Deidentified results and analysis outputs may be available upon reasonable request.

## Code Availability

The analysis and deployment code will be publicly available upon publication at https://github.com/groeschel-lab/inconsistency-check

## Notes

### Competing Interest Statement

Markus Vogel is employee of Microsoft Germany GmbH

### Author Declarations

Ethics committee/IRB of Charite University Medicine Berlin gave ethical approval for this work (EA2/322/23)

## References

1. Williams CYK, Subramanian CR, Ali SS, et al. Physician- and large language model-generated hospital discharge summaries. JAMA Intern Med 2025;185(7):818–25.

2. Shool S, Adimi S, Saboori Amleshi R, Bitaraf E, Golpira R, Tara M. A systematic review of large language model (LLM) evaluations in clinical medicine. BMC Med Inform Decis Mak 2025;25(1):117.

3. Jung K-H. Large language models in medicine: Clinical applications, technical challenges, and ethical considerations. Healthc Inform Res 2025;31(2):114–24.

4. Hameed BMZ, Naik N, Ibrahim S, et al. Breaking barriers: Unveiling factors influencing the adoption of artificial intelligence by healthcare providers. Big Data Cogn Comput 2023;7(2):105.

5. Montagnese M, Rangelov B, Doel T, et al. Cloud computing for equitable, data-driven dementia medicine. Lancet Digit Health 2025;7(11):100902.

6. Guerriero M, Garriga M, Tamburri DA, Palomba F. Adoption, support, and challenges of infrastructure-as-code: Insights from industry [Internet]. In: 2019 IEEE International Conference on Software Maintenance and Evolution (ICSME). IEEE; 2019. DOI: 10.1109/icsme.2019.00092

7. Beaulieu-Jones BK, Greene CS. Reproducibility of computational workflows is automated using continuous analysis. Nat Biotechnol 2017;35(4):342–6.

8. Haynes AB, Weiser TG, Berry WR, et al. A surgical safety checklist to reduce morbidity and mortality in a global population. N Engl J Med 2009;360(5):491–9.

9. Boussina A, Krishnamoorthy R, Quintero K, et al. Large language models for more efficient reporting of hospital quality measures. NEJM AI [Internet] 2024;1(11). DOI: 10.1056/aics2400420

10. Hirsch A, Ten FW, Krüger KS, et al. The Multimodal Anonymizer: a fully local multi-agent AI system for medical data deidentification [Internet]. medRxiv. 2026;DOI: 10.64898/2026.05.28.26353952

11. Ancker JS, Edwards A, Nosal S, et al. Effects of workload, work complexity, and repeated alerts on alert fatigue in a clinical decision support system. BMC Med Inform Decis Mak 2017;17(1):36.

12. Liang AS, Amrollahi F, Jiang Y, et al. SmartAlert - implementing machine learning-driven clinical decision support for inpatient laboratory utilization reduction. NEJM AI [Internet] 2026;3(7). DOI: 10.1056/aics2501086

