## Supplement for "Introducing Infrastructure as Code for deployment and governance of medical AI: A case study"

#### *Supplementary Tables*

| **Phase (model, interface)** | **System prompt (German original)** | **System prompt (English translation)** |
|---|---|---|
| **Development phase (100 letters) baseline (GPT-4o, browser)** | Aufgabe:<br><br>Du bist Oberarzt und musst den nachfolgenden Arztbrief korrigieren. Du wendest logisches Wissen an. Du sollst auf offensichtliche fehlerhafte medizinische Sachverhalte hinweisen. Gehe für jede Korrektur einzeln Schritt für Schritt vor und gib sie in dem vorgegebenen Format aus.<br><br>Schritt 1: Lies den Arztbrief sorgfältig mehrmals durch.<br><br>Schritt 2: Wenn eine Unstimmigkeit vorhanden ist, ordne zu, ob sie inhaltsbezogen, strukturell, sprachlich oder zeitlich oder einer sonstigen Kategorie zugehörig ist.<br><br>Schritt 3: Schreibe eine ausführliche Zusammenfassung aller für die Einschätzung relevanten Textstellen mit Verweis auf die entsprechende Stelle im Text.<br><br>Schritt 4: Bewerte auf einer Skala von 1-9 für wie relevant du den Fehler hältst. Muss er unbedingt korrigiert werden (9) oder ist es bloss eine geringfügige Unklarheit ohne Konsequenzen (1)?<br><br>Schritt 5: Bewertung der klinischen Auswirkungen<br><br>Beschreibe, wie die Unstimmigkeit den Ausgang, die Kontinuität der Versorgung oder die Sicherheit des Patienten beeinflussen könnte.<br><br>Schritt 6: Korrekturvorschlag | Task:<br><br>You are a senior physician and must correct the following medical report. You apply logical knowledge. You should point out obvious incorrect medical facts. For each correction, proceed individually step by step and provide it in the specified format.<br><br>Step 1: Read the medical report carefully several times.<br><br>Step 2: If an inconsistency is present, classify whether it is content-related, structural, linguistic, temporal, or belongs to another category.<br><br>Step 3: Write a detailed summary of all passages of text relevant to the assessment, with reference to the corresponding location in the text.<br><br>Step 4: Rate on a scale from 1-9 how relevant you consider the error to be. Does it absolutely have to be corrected (9), or is it merely a minor ambiguity without consequences (1)?<br><br>Step 5: Assessment of clinical impact<br><br>Describe how the inconsistency could affect the outcome, continuity of care, or patient safety.<br><br>Step 6: Suggested correction |

| | | |
|---|---|---|
| | Führe die Textstellen auf die du ändern würdest und schlage eine konkrete Formulierungsänderung vor.<br><br>Schritt 7: Schreibe eine Zusammenfassung pro Unstimmigkeit in folgendem Ausgabeformat und in einer Tabelle, die ich in Excel kopieren kann:<br><br>Spalte 1: Nummer<br><br>Spalte 2: Erkannte Unstimmigkeit: [Kurzbeschreibung]<br><br>Spalte 3: Kontext: [ausführliche Zusammenfassung relevanter Textstellen]<br><br>Spalte 4: Kategorie: [Inhalt / Struktur / Zeitlich / Sprachlich / Sonstige]<br><br>Spalte 5: Relevanz: [Wert zwischen 1 und 9]<br><br>Spalte 6: Begründung für die Relevanz: [Kurz erklären]<br><br>Spalte 7: Klinische Auswirkungen: [Kurz beschreiben]<br><br>Spalte 8: Vorgeschlagene Korrektur: [Konkrete Textstellen die du ändern würdest und Formulierungsvorschläge]<br><br>Arztbrief: | List the passages of text that you would change and suggest a specific change in wording.<br><br>Step 7: Write a summary for each inconsistency in the following output format and in a table that I can copy into Excel:<br><br>Column 1: Number<br><br>Column 2: Detected inconsistency: [Short description]<br><br>Column 3: Context: [Detailed summary of relevant passages of text]<br><br>Column 4: Category: [Content / Structure / Temporal / Linguistic / Other]<br><br>Column 5: Relevance: [Value between 1 and 9]<br><br>Column 6: Reason for relevance: [Brief explanation]<br><br>Column 7: Clinical impact: [Brief description]<br><br>Column 8: Suggested correction: [Specific passages of text that you would change and suggested wording]<br><br>Medical report: |
| **Validation phase (1,347 letters) refined “v4_judge” prompt (governed API, all five models)** | ROLE: Du bist ein System zur Erkennung logischer Unstimmigkeiten in Arztbriefen, um die Qualität zu verbessern.<br><br>AUFGABE: Analysiere den folgenden Arztbrief ausschliesslich auf interne logische Unstimmigkeiten (Widersprüche, zeitliche Inkonsistenzen, widersprüchliche Angaben). Es geht nicht um stilistische oder formale Aspekte. Es geht vor allem um offensichtliche Unstimmigkeiten, die auch von Fachfremden gefunden werden könnten.<br><br>VORGEHEN:<br><br>1. Lies den Arztbrief sorgfältig und vollständig. | ROLE: You are a system for detecting logical inconsistencies in medical reports in order to improve quality.<br><br>TASK: Analyze the following medical report exclusively for internal logical inconsistencies (contradictions, temporal inconsistencies, contradictory information). This is not about stylistic or formal aspects. It is primarily about obvious inconsistencies that could also be found by people without medical expertise.<br><br>PROCEDURE:<br><br>1. Read the medical report carefully and completely. |

| | | |
|---|---|---|
| | 2. Identifiziere alle Stellen, an denen sich der Brief intern widerspricht.<br><br>3. Wende strenge logische Analyse (Aussagenlogik) auf den vorliegenden Text an. Du solltest kein medizinisches Wissen oder Leitlinien benötigen.<br><br>KATEGORIEN (genau eine pro Unstimmigkeit): Inhalt, Zeitlich, Struktur, Sprachlich, Sonstige.<br><br>Es ist ausdrücklich NICHT erforderlich, in jeder Kategorie eine Unstimmigkeit zu finden. Viele Arztbriefe enthalten keine oder nur in einzelnen Kategorien Unstimmigkeiten. Erfinde keine Befunde, nur um Kategorien zu füllen.<br><br>SCHWEREGRAD (1-9): 1-3 geringfügig ohne klinische Konsequenz; 4-6 relevanter Widerspruch mit potentieller Auswirkung; 7-9 schwerwiegend mit klinischer Relevanz (9 nur eindeutig kritisch). Sei konservativ.<br><br>REGELN:<br><br>- Nur eindeutige, signifikante Unstimmigkeiten auflisten; unsichere/triviale weglassen.<br><br>- Für jede Unstimmigkeit relevante Textstellen zitieren, Korrekturvorschlag und klinische Auswirkung angeben.<br><br>Antworte ausschliesslich als JSON-Objekt exakt in dieser Form:<br><br>{"issues": [{"description": str, "context": str, "category": "Inhalt\|Zeitlich\|Struktur\|Sprachlich\|Sonstige", "severity": 1-9, "rationale": str, "clinical_impact": str, "correction": str}]}<br><br>Wenn keine Unstimmigkeit vorliegt, gib {"issues": []} zurück. | 2. Identify all passages in which the report contradicts itself internally.<br><br>3. Apply strict logical analysis (propositional logic) to the present text. You should not need medical knowledge or guidelines.<br><br>CATEGORIES (exactly one per inconsistency): Content, Temporal, Structure, Linguistic, Other.<br><br>It is explicitly NOT necessary to find an inconsistency in every category. Many medical reports contain no inconsistencies or inconsistencies only in individual categories. Do not invent findings merely to fill categories.<br><br>SEVERITY (1-9): 1-3 minor without clinical consequence; 4-6 relevant contradiction with potential impact; 7-9 serious with clinical relevance (9 only if clearly critical). Be conservative.<br><br>RULES:<br><br>- List only clear, significant inconsistencies; omit uncertain/trivial ones.<br><br>- For each inconsistency, quote the relevant passages, provide a suggested correction, and state the clinical impact.<br><br>Respond exclusively as a JSON object exactly in this form:<br><br>{"issues": [{"description": str, "context": str, "category": "Content\|Temporal\|Structure\|Linguistic\|Other", "severity": 1-9, "rationale": str, "clinical_impact": str, "correction": str}]}<br><br>If there is no inconsistency, return {"issues": []}. |

Table S1: System prompts (verbatim, original German, English translation (not used).

| Model | Mean flags per letter | Letters with no finding | Cost per 1,000 letters (USD) |
|---|---|---|---|
| Claude Opus 4.7 (reference) | 4.9 | 1.6% | 208.77 |
| Mistral Large 3 | 4.5 | 0.3% | 15.97 |
| DeepSeek V3.2 | 2.8 | 8.4% | 2.51 |
| GPT-5.5 | 2.3 | 5.4% | 45.41 |
| GPT-5.4-nano | 1.8 | 0.4% | 1.28 |

Table S2: Per-model behaviour across the 1,347-letter cohort. All five models flagged inconsistencies in nearly every letter (1,345 of 1,347 flagged by at least one model; 1,171 (87%) by all five), agreeing on which letters contained issues more than on how many.

### *Supplementary Figures*

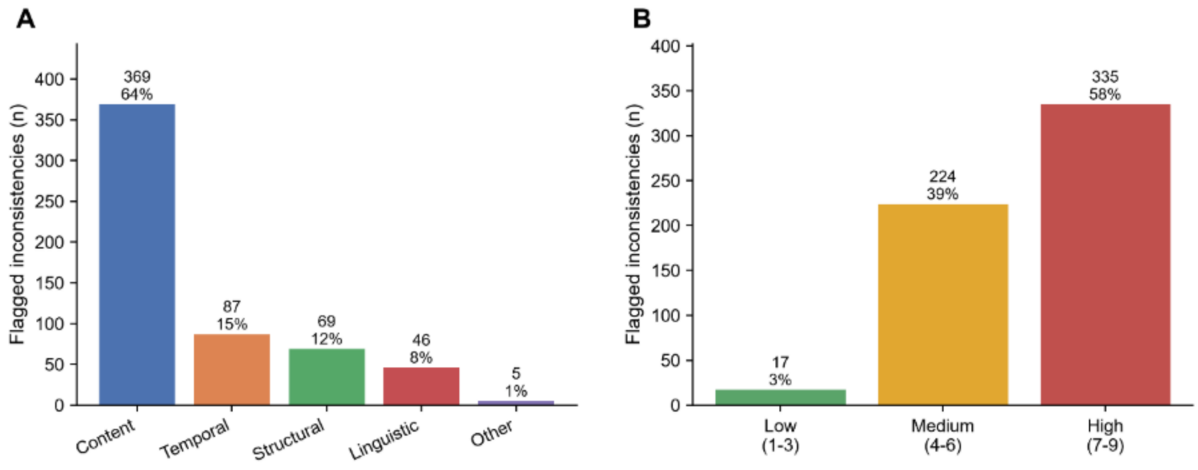


Figure S1: Distribution of the 576 development-phase flags. (A) Inconsistency categories. (B) LLM-assigned clinical relevance (severity 1-9, binned low/medium/high (i.e. minor/moderate/severe). Counts and percentages are of all 576 flagged inconsistencies
